# Optimising access to vocational rehabilitation through multiple sclerosis charities: Protocol for a feasibility randomised controlled trial

**DOI:** 10.1101/2025.05.18.25327869

**Authors:** Blanca De Dios Perez, Denise Kendrick, Roshan das Nair, Nikos Evangelou, Ian Newsome, Kate Radford

**Affiliations:** University of Nottingham (Centre for Rehabilitation and Ageing Research, Queens Medical Centre), Nottingham, UK; NIHR Nottingham Biomedical Research Centre, Nottingham, UK; Centre for Academic Primary Care, Lifespan and Population Health, School of Medicine, University of Nottingham, Nottingham, UK; University of Nottingham (Mental Health & Clinical Neurosciences, School of Medicine); Nottinghamshire Healthcare Trust (Institute of Mental Health); SINTEF (Health Division), Trondheim, Norway; Lay co-author, York, UK

**Keywords:** Vocational rehabilitation, multiple sclerosis, charities, employment support, job retention

## Abstract

**Background:** People with multiple sclerosis (MS) often leave the workforce prematurely due to MS symptoms and difficulties managing workplace relationships and performance. Vocational rehabilitation (VR) can improve job retention outcomes for people with MS, but there is a lack of evidence on the effectiveness of these interventions.

**Methods:** A multicentre, feasibility, parallel-group randomised controlled trial (RCT) comparing a job retention VR intervention plus usual care (n=30) with usual care alone (n=30). This study includes an embedded mixed-methods process evaluation. People with MS, aged 18-65 years, in paid employment will be recruited from MS charities.

Participants with MS will be able to include their employers in the intervention to receive information about MS and employment. The intervention involves an initial interview and up to 10 hours of employment support for people with MS and up to four hours of support for employers, over six months. Employees from MS charities will be recruited and trained to deliver the MSVR intervention. Participants will be followed up by postal/telephone/online questionnaires at 6-, 9-, and 12-months post-randomisation.

The aim is to ascertain the feasibility and acceptability of delivering the intervention within MS charities, and to determine parameters for future trial and explore the acceptability of the study intervention and procedures.

**Discussion:** This novel study will provide insight into how existing services from MS charities can fill a service gap by providing employment support to people with MS. Findings will inform the design of a future fully powered RCT.

**Trial Registration Number:** NCT06966115

## Introduction

Multiple sclerosis (MS) is a complex health condition affecting the central nervous system (CNS) that leads to a range of physical, emotional, and cognitive difficulties [1,2].

MS can significantly impact employment outcomes [3], with a high number of people with MS leaving the workforce a few years post-diagnosis due to symptoms such as fatigue, cognitive difficulties, mobility limitations, and mood difficulties [4–6]. In fact, fatigue is one of the main factors leading to individuals leaving the workforce [7].

Vocational rehabilitation (VR) can enable people with illnesses or disabilities to remain in paid employment by modifying the environment through reasonable adjustments and providing strategies to manage symptoms that negatively impact work performance [8]. The VR needs of people with MS vary according to the complexity of their circumstances, with some only needing information provision and others needing multidisciplinary support [9]. Who needs what level of support based on employment, clinical, and demographic characteristics remains unclear.

VR for people with MS is varied and could involve a range of activities to support symptom management and modify the work environment. For example, reasonable adjustments (e.g., modifications to the work environment or duties) can help people with MS manage their symptoms and remain at work longer [10,11]. However, research indicates that individuals with MS often require support and encouragement to obtain reasonable adjustments in the workplace through effective negotiation with their employer or organisation [12]. Unfortunately, people with MS and employers do not always understand the support that would be beneficial at work and have limited access to services providing such advice [13,14].

We have previously developed a psychologist and occupational therapist-delivered VR intervention to support people with MS to remain in paid employment [15]. We have tested this programme in a community setting [13] and within the UK National Health Service (NHS) [16]. The preliminary findings demonstrate that the intervention is acceptable to people with MS and their employers and helps participants with MS to meet their vocational goals [13,16]. However, we also identified some barriers (e.g., lack of staff, service structure, etc.) to deliver this programme more widely within a healthcare setting [16,17]. In view of this, patient and public involvement (PPI) representatives suggested testing our VR intervention within the services provided by MS charities. This approach could be beneficial to reach people with MS who might be in the earlier stages of the illness and looking for information or who have not considered discussing “work” with their healthcare providers [17].

We have therefore adapted our VR intervention, called MSVR (“multiple sclerosis vocational rehabilitation”), to be delivered by non-specialists working for MS charities. The practice of training non-specialists to deliver evidence-based and complex interventions has increased over recent years to reach communities where there is a lack of resources or to help increase the reach of these interventions [18,19].

Before conducting a fully powered randomised controlled trial (RCT), the feasibility of delivering our adapted VR intervention through MS charities needs to be established. This study was informed by the Medical Research Council (MRC) guidance for developing and evaluating complex interventions [20].

### Aims and objectives

The aim is to assess the feasibility of conducting a multicentre, parallel-group RCT comparing MSVR plus usual care (UC) delivered to participants recruited from MS charities with UC alone. UC comprises the existing services offered by the charities. The objectives are:

#### 1. Recruitment

To assess how long it takes to recruit the planned sample, the recruitment rate, the proportion of eligible people with MS recruited, reasons for non-recruitment, and the suitability of study characteristics (e.g., inclusion and exclusion criteria).

#### 2. Retention and engagement

To assess the proportion of participants lost to follow-up and the reasons for loss to follow-up, withdrawal rates, data completeness, participants’ compliance with MSVR, and identify factors affecting compliance.

#### 3. Intervention delivery

To assess the feasibility of delivering up to 10 hours of VR per participant over 6 months, intervention fidelity, intervention adherence, feasibility of delivering VR alongside the charities’ existing services, and practical issues related to intervention delivery.

#### 4. Acceptability

To explore through an embedded mixed-methods process evaluation the acceptability of MSVR and study procedures for participants with MS, employers, and employees from MS charities delivering the intervention (hereafter referred to as MSVR champions).

#### 5. Trial parameters

To determine parameters for calculating the sample size for a future large-scale RCT.

#### 6. Training of staff

To explore the mentoring and training needs of the employees from MSVR champions recruited for the study.

#### 7. Study resources

To explore the usefulness of the MSVR training manual, training sessions and mentoring support during the trial.

#### 8. Intervention cost

To estimate the cost of training MSVR champions and delivering MSVR to people with MS and their employers.

## Methods and Analysis

### Study Design

Multicentre, feasibility, parallel-group RCT with an embedded mixed methods process evaluation. Participants will be randomised (1:1 ratio) to receive either MSVR plus UC or UC alone. The intervention will be delivered remotely in collaboration with MS charities in the United Kingdom (UK).

This protocol conforms to the SPIRIT (standard protocol items: recommendations for interventional trials) guidelines [21] (see S1). The findings from this feasibility randomised controlled trial (fRCT) will be reported following the CONSORT 2010 statement: extension to randomised pilot and feasibility trials [22].

### Setting and locations

The University of Nottingham will coordinate this study, involving up to six MS charities. The charities will be purposively recruited based on their attributes (e.g., organisation size, resources, and services available) and interest in providing the VR intervention. The intervention will be delivered remotely via Microsoft Teams and/or telephone, according to the participants’ preferences. The study will be conducted between June 2025 and November 2027.

### Eligibility criteria

People with MS (1) aged ≥18, who can (2) give informed consent, (3) communicate in English, and (4) who are in paid employment (including self-employed). This study will exclude people planning to retire within the next 12 months or receiving VR support from the NHS or other employer services.

If a participant with MS consents to involving their employer in the intervention, they will be included if they are (1) aged ≥18, who can (2) able to give informed consent,(3) communicate in English, and (4) currently employing a person with MS. We will consider the line manager or a human resources representative as the employer of the person with MS. There are no exclusion criteria for the employers.

MSVR champions recruited to deliver the intervention will be asked to consent to complete a questionnaire following the training sessions and to participate in an interview at the end of the study.

### Recruitment

Participants with MS will be recruited through the participating MS charities over a 10-month period. Recruitment will start in June 2025 and conclude in March 2026. The organisations will share information about the study using a multi-pronged approach to reach a broad range of people with MS in terms of clinical, employment and demographic characteristics. Therefore, the study will be advertised on their helplines, social media, websites, webinars, and bulletins.

The initial approach to potential participants with MS will be made by the MS charities’ employees through their helpline support, newsletters, and events. They will provide information about the study using a study advert and participant information sheet (PIS). People with MS declining to participate will be asked for a reason for not participating.

People with MS will have the opportunity to read about the study online (e.g., social media, websites, bulletins). If a person with MS is interested in participating, they will be asked to complete an expression of interest with the charity worker to consent for their details to be shared with the researcher.

Those people with MS who are interested in participating in the study will be told that they can include their employer (e.g., line manager or human resources representative) in the intervention to receive information and advice about MS at work. It is not mandatory for the person with MS to include their employer, and if the person with MS does not consent to include their employer, the employer will not be contacted.

### Screening and informed consent

Potentially eligible participants with MS will be contacted via telephone to be screened against the inclusion criteria by the lead researcher. A screening log will be used to monitor and record information about participants screened and the reasons for not being recruited.

Following screening, potential participants who want to participate will complete a written consent form. Potential participants will receive an email with a link to an electronic consent form from REDCap. Those who prefer not to use REDCap to complete the consent form and baseline questionnaire via the online link will be able to do so by completing the written consent form verbally over the telephone or via video conference with a research team member. All participants will receive a copy of their signed and completed consent form via email.

### Intervention

#### MSVR

MSVR will be delivered by employees working within MS charities supporting people with MS with advocacy or information provision. These employees have a wide range of professional backgrounds, expertise in MS (including symptoms and progression), skills with active listening, and have been trained to handle difficult conversations. The workers from MS charities, called “MSVR Champions”, will receive training and mentoring in preparation for the trial.

MSVR for people with MS involves an initial assessment (including vocational goal setting), followed by up to 10 hours of individually tailored support according to need, for up to 6 months. The support will focus on helping the person with MS to meet their vocational goals by addressing topics such as:

- Understanding MS
- Advice on reasonable adjustments (e.g., modification to the work environment or duties to accommodate the impact of MS)
- Support requesting reasonable adjustments
- Fatigue management
- Managing cognition at work
- Information about legal rights
- Disclosure (i.e.., telling the employer or colleagues about the MS diagnosis)
- Long-term career planning (i.e., exploring alternative career options)
- Managing mood difficulties
- Signposting to local and national resources.

To improve intervention adherence, the intervention will be delivered remotely (e.g., via telephone, videoconference, or email) to reduce the impact of fatigue on travelling and preparing for appointments. The intervention also allows great flexibility in arranging the sessions at a time convenient for the person with MS. The topics addressed will vary according to the complexity of their workplace difficulties. Participants will receive a brief written summary of the content discussed in each session via email, with further resources to complement the information provided.

MSVR for the employers will involve an initial interview (approximately 30 minutes) to understand their experiences supporting the employee with MS and up to 4 hours of support for up to six months. The employers’ intervention includes:

- Signposting to relevant organisations.
- Educational resources.
- Information about MS and invisible symptoms.
- Support with identifying reasonable adjustments.
- Legal responsibilities (Equality Act).

After each session, the employer will receive an email summarising topics discussed and, when required, a list of actions they should complete before the next session.

### Control Condition

Participants (MS and employers) in the control group will receive existing services available from the MS charities’ websites, helplines, and events. Due to the nature of the study, UC will vary between participants. However, no MS charity recruited for this study will have a specialist VR service (and none should have indicated that they will be creating such a service within the study time frame).

Participants (MS and employers) in the control group will interact with members of the MS charity who have not been trained and are not involved in MSVR delivery to reduce the potential for contamination.

UC was chosen as the comparator intervention to establish a benchmark based on existing services, both for ethical considerations and to ensure that participants continued to have access to these support services.

At the end of the study, participants with MS in the control group will receive the “Work and MS: An Employee’s Guide” booklet from the MS Society, which contains information about MS and work. PPI representatives suggested this to reduce the burden of being allocated to the control group and encourage the completion of follow-up questionnaires.

#### Concomitant Therapy

Participants will continue to use services from social care, the NHS, and other third- sector organisations alongside MSVR. Information on any concomitant therapy received will be collected using a resource use questionnaire.

### Monitoring and Mentoring

An Occupational Therapist (OT) with extensive experience in VR for people with long-term health conditions will offer monthly mentoring sessions to the MSVR champions.

The mentoring sessions will be conducted in groups via videoconference calls to discuss cases, confidentiality issues, contamination, and challenges during the intervention delivery. MSVR champions will also be able to request further mentoring if a query arises outside the mentoring sessions.

The OT will complete a mentoring record form to capture topics addressed, challenges related to MSVR delivery, and questions raised by MSVR champions during mentoring to gain further understanding of the skills needed to deliver MSVR and additional training needs.

### Outcomes

The first outcomes assess the feasibility and acceptability of delivering MSVR within the context of MS charities by measuring recruitment, retention, engagement, and intervention delivery.

Secondary outcomes refer to determining the parameters for a fully powered trial, estimating sample size, intervention costs, and identifying intervention improvements and further training needs of MSVR champions. The feasibility objectives are summarised in Table 1.

**Table 1:**
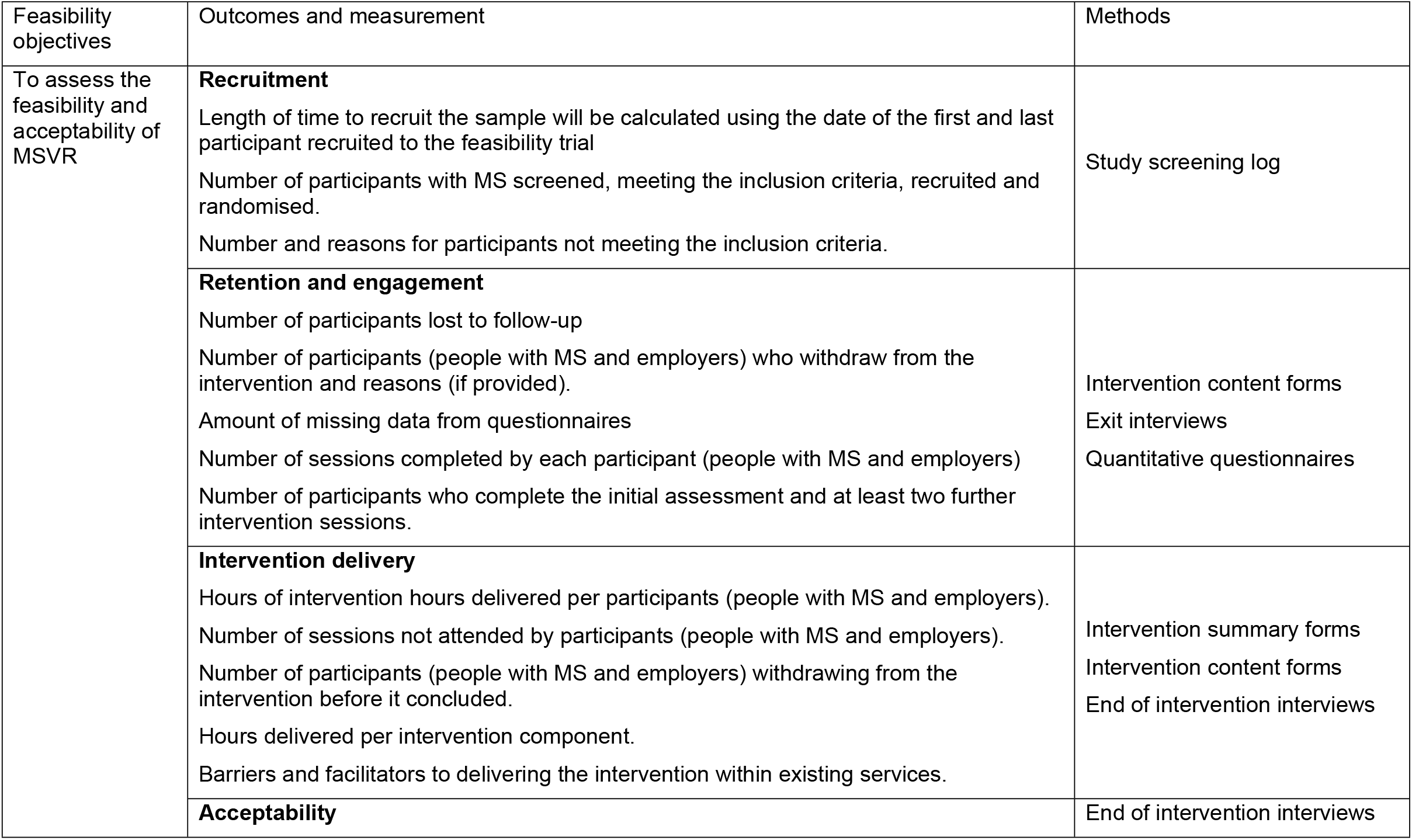
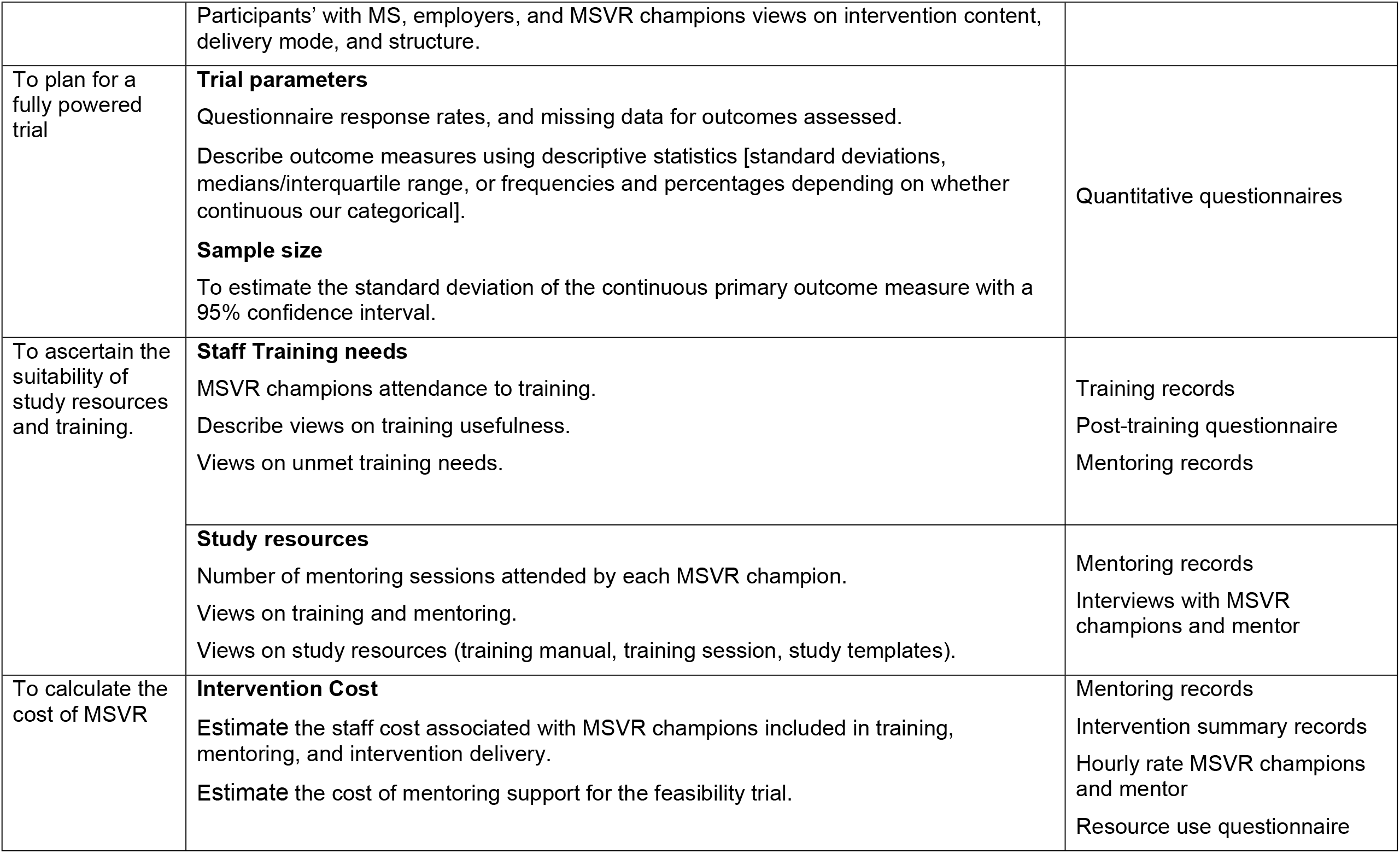
Feasibility objectives, measures, and methods.

### Outcome measures

Outcome measures will be assessed at 6-, 9-, and 12-months post-randomisation, with data collection completion expected in March 2027. The measures collected and time points have been summarised in Fig 1.

**Figure 1.**
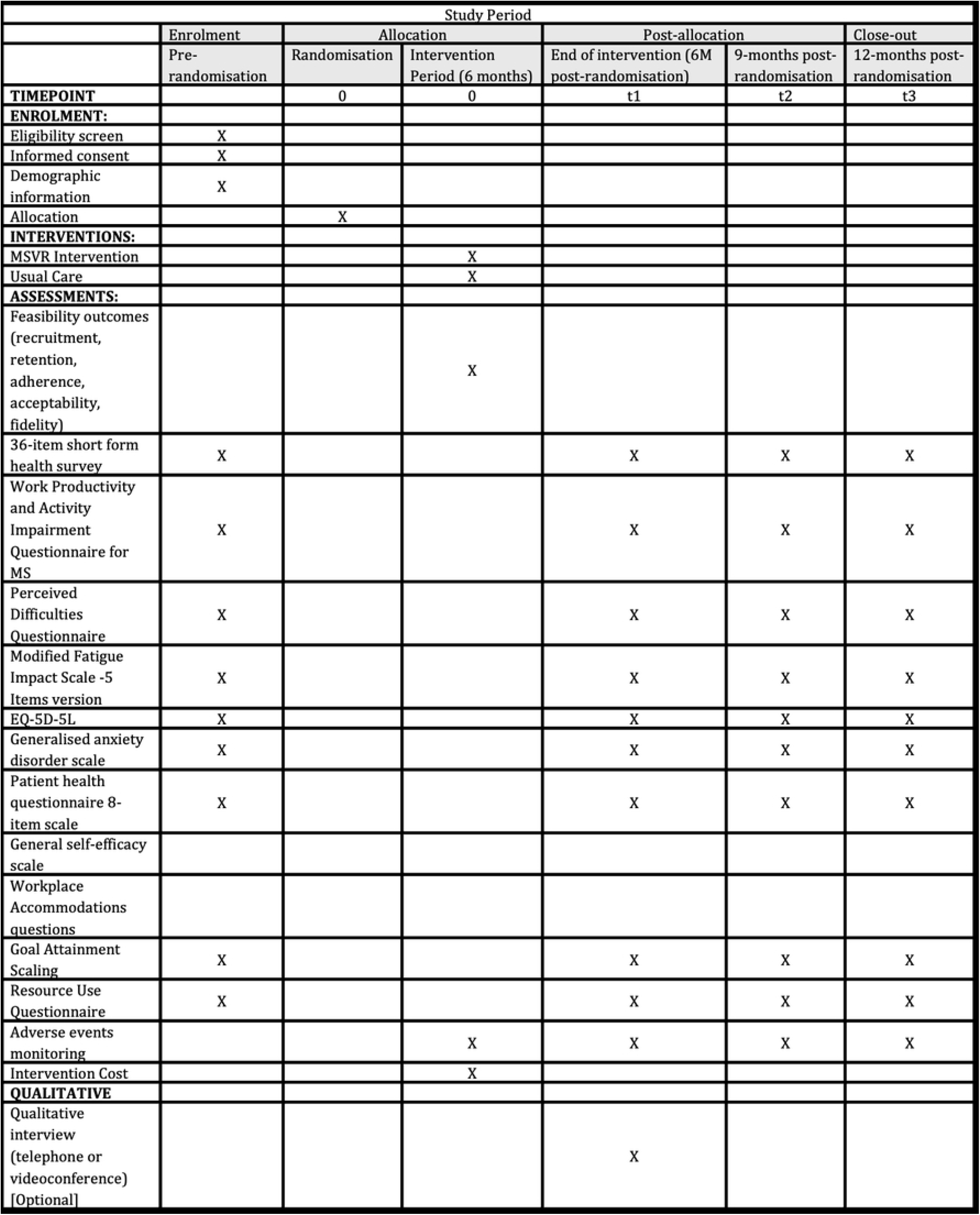
SPIRIT Figure — Feasibility Trial Schedule of Enrolment, Interventions, and Assessment

The outcome measures will be collected using REDCap, or by post or telephone if a participant cannot access an electronic device. The acceptability measures (i.e., interviews) will be collected remotely via telephone or Microsoft Teams. To minimise missing data, we will set up a REDCap email notification to automatically remind participants to complete the questionnaires 2 weeks from the due date. Additional approaches to reduce missing data include reminders via text message, phone and postal reminders.

Participants completing the questionnaire by post or telephone will be contacted via email or telephone to remind them to complete the questionnaire. If needed, a second copy of the booklet of questionnaires will be sent by post. Participants will receive priming calls to remind them to complete the questionnaires.

To improve the rate of questionnaire completion, participants with MS and employers will be offered a £20 retail voucher if they complete all questionnaires.

### Outcome measures

#### Participants with MS

This feasibility study has two candidate coprimary outcome measures for a future definitive trial: (1) health-related quality of life as measured by the 36-item short form health survey (SF-36) [23] and (2) work productivity, measured using the Work Productivity and Activity Impairment Questionnaire (WPAI) for people with MS [24], extending the recall period from seven days to four weeks, which has been shown to be a reliable measure. These measures have been identified as relevant in other studies about MS and employment [25,26].

The SF-36 measures eight health constructs relating to physical and mental health. This measure has a high internal consistency (Cronbach’s alpha >0.80) and good test-retest reliability (>0.70). The WPAI:MS measures absenteeism and presenteeism, productivity loss and activity impairment. Higher scores in the WAIP:MS represent a higher impact of MS at work and in conducting everyday activities.

The European Quality of Life with five dimensions (ED-5D-5L) [27] will assess psychological and health-related quality of life, focusing on mobility, self-care, usual activities, pain/discomfort, and anxiety/ depression. Cognitive difficulties will be measured with the perceived deficits questionnaire (PDQ) [28,29], a 5-point Likert scale 20-item questionnaire with a high internal consistency (Cronbach’s alpha .93). Fatigue will be assessed using the modified fatigue impact scale 5 (MIFS-5), a 4- point Likert scale with five items [30]. The General Self-Efficacy Scale (GSES) assesses the participants’ belief in their ability to complete tasks successfully [31]. Two brief questionnaires will assess mood: the Generalised Anxiety Disorder Scale (GAD-7) [32] and the Patient Health Questionnaire (PHQ-8) [33].

Vocational goals will be measured using the goal attainment scale (GAS) [34] to ascertain the impact of the intervention on the vocational goals set by participants at the beginning of the study. Workplace accommodations will be assessed using a series of binary questions (yes/no) about support received at work. These questions were extracted from a study reporting workplace accommodations for stroke survivors [35].

Finally, a resource use questionnaire will be developed to record healthcare utilisation and additional services used during the feasibility trial.

#### Employers

Employers will be asked to complete three questions, developed for this study, regarding their knowledge of MS, confidence in managing the employee with MS at work, and confidence in their ability to solve future problems at work.

#### MSVR Champions

MSVR champions will be asked to complete the educational course assessment toolkit (EDUCATOOL) [36]. A 12-item questionnaire will be used to evaluate the training and learning experience of the MSVR champions during the study.

### Progression Criteria to the Future Trial

Progression criteria will be assessed based on recruitment and follow-up rates based on a traffic light system of green (go), amber (review), and red (stop) [37]. The start of recruitment is defined as the first participant recruited into the trial.

Recruitment criteria will be assessed after 3-4 months to allow recruitment rates to stabilise. The follow-up criteria will be assessed at the end of the intervention.

Feasibility will be demonstrated if:

- At least six people with MS are recruited per month across all sites (green), at least three but less than six (amber), or less than three (red).
- Percentage of participants randomised to the MSVR arm who complete the intervention (initial assessment and at least two sessions), at least 80% green, at least 40% but less than 80% (amber) or less than 40% (red).
- The follow-up rate criterion is assessed by the percentage of people who complete the end of intervention questionnaire with at least 80% (green), at least 65% but less than 80% (amber) or less than 65% (red).

A rescue plan has been developed to ensure the progression criteria are met. If the rescue plan is not effective (e.g., progression criteria measures do not progress to amber or green), and any criteria are graded as red, we will not progress to the definitive trial. The findings from the progression criteria stages throughout the trial will be used to inform the full-scale future trial.

### Sample size

A pragmatic target of 60 participants with MS (30 in each arm) was estimated based on personal communications with the MS charities recruited for the study and experiences from previous studies [13,16]. This figure aligns with recommended sample sizes for feasibility trials to estimate a parameter for a future RCT [38–40]. This sample size will provide sufficient data to achieve the study aims mentioned above.

Participants with MS will be asked if they are interested in recruiting their employer to the intervention, where they will receive information and advice about MS and employment. Based on previous research studies [13,16], we anticipate that approximately 20% of participants will consent to include their employer.

### Randomisation

Following the completion of the consent form, participants will have the opportunity to complete the baseline assessment electronically (using REDCap) or over the telephone with a member of the research team.

After completing the consent form and baseline assessment, MS participants will be automatically randomised (1:1 ratio) to MSVR plus UC or UC, using adaptive randomisation with minimisation through a computer. The balancing variables selected for the minimisation are the study site and the type of MS, classified as progressive or non-progressive MS. The MSVR champions will be notified by email, informing them of the group allocation.

### Blinding

Due to the nature of the intervention, the MSVR champions and the mentor will not be blinded to the intervention group of the participants. Participants will not be blinded to the intervention group allocation. The teams at the MS charities will not be able to predict the allocation group when referring participants to the study.

Follow-up data will be collected, wherever possible, using REDCap. For participants who prefer to complete the questionnaire via post or telephone, a member of the research team, blinded to allocation group will collect the data. A form will be developed to record whether the researcher was unblinded during the data collection process.

The researcher supporting the recruitment of participants and conducting the data analysis will be blinded to the group allocation. Other research team members not involved in the direct management of the feasibility trial will remain blinded to group allocation until data collection concludes.

Unblinding of treatment allocation will occur after the final participant has completed the 9-month post-randomisation follow-up questionnaire and data have been entered and validated.

### Intervention adherence and fidelity measures

Intervention adherence will be measured using the intervention summary form, collecting data on sessions attended, missed, reasons (if provided), and intervention length.

Intervention fidelity will be measured using the “intervention content forms”. MSVR champions will be trained to complete these forms at the end of each session, reporting the modality of the session, topics discussed, length of time spent discussing each topic, and time spent by MSVR champion on non face-to-face activities after the session (e.g., liaison with other professionals, reviewing forms, etc).

An intervention fidelity checklist will monitor whether the MSVR champions deliver the intervention as intended. To explore this, MSVR champions will be asked to video or audio record at least 10% of the intervention sessions. A research assistant will analyse the session’s content compared to the data recorded in the “session content form” to identify discrepancies.

### Reporting and management of adverse events

Serious Adverse Events (SAE) involve any untoward occurrence that results in death, is life-threatening, requires hospitalisation or prolongation of existing hospitalisation, results in persistent or significant disability or incapacity, or is otherwise considered medically significant by the investigator. Due to the nature of the intervention, we do not anticipate SAEs.

Related unexpected serious adverse events (RUSAE) will be any SAE where, in the opinion of the Principal Investigator (PI), the event was considered to be (1) “related” that is, it resulted from the administration of any of the research procedures, and (2) “unexpected” that is, the type of event is not listed in the protocol as an expected occurrence. In the case of this study, it could involve accidental injury resulting from workplace adaptations recommended by the MSVR champion, or workplace accidents resulting in injury requiring hospital treatment.

This data will be collected by self-report (participant questionnaires and ad-hoc CRFs, or via sites notifying a member of the research team). Any event that, in the opinion of the PI, is “related” (resulted from the administration of the research procedure) and “unexpected” (the type of event is not listed as an expected occurrence) will be reported to the research ethics committee within 7 days of being informed of such events. Fatal and life-threatening events will be reported no later than 3 calendar days after the sponsor or PI is first aware of the event. Any other additional relevant information will be reported within 7 calendar days of the initial report.

### Process Evaluation

An embedded process evaluation will follow the MRC guidelines for process evaluation of complex interventions [41]. This process will be informed by normalisation process theory [42] to understand how the intervention interacted with the context of the charities recruited for the study and ascertain how the intervention needs to be adapted to achieve long-term sustainability within the new context.

To refine the study procedures and intervention for a future trial, we will seek the views of participants with MS (in the intervention and control groups), employers, and MSVR champions. We will use purposive sampling to recruit participants with diverse employment characteristics. We will collect data on participants who dropped out and participants who did not book intervention sessions to explore the characteristics between those who completed the intervention and those who did not. A research team member will conduct the interviews via telephone or video conference (e.g., Microsoft Teams).

At the end of the intervention, semi-structured interviews with at least 21 participants [participants with MS (n=10), employers (n=5), MSVR champions (n=5), and OT mentor (n=1)] will be conducted to explore the acceptability of MSVR, factors affecting their engagement in the intervention, acceptability of the intervention, their views on what attributes of the intervention where most beneficial for them, explore whether the intervention needs to be further adapted for delivery within MS charities, and views on how to improve the intervention for a future larger trial. Participants with MS will be asked what can be done in a future trial to encourage employer recruitment during the semi-structured interviews.

The MSVR champion interviews will also explore the impact of mentoring on intervention delivery. Interviews with control group participants will also examine whether participants actively sought support with employment elsewhere and whether they took self-directed action towards their goals.

### Trial management

BDP has the overall responsibility for the feasibility trial and will receive support from the co-investigators (KR, DK, RdN, NE) to manage and coordinate the study. An independent steering committee, including academics and PPI representatives, will meet every two months to discuss progress.

Due to the nature of the study (i.e., feasibility), a data monitoring committee will not be needed.

## Data analysis

The study’s quantitative data will be analysed using SPSS version 27.0 (Statistical Package for Social Sciences) following a predefined statistical analysis plan. Data from REDCap will be extracted onto an Excel document and stored using OneDrive encrypted storage from the University of Nottingham. Participants will be given a participant ID to maintain anonymity of the data, and personal data will be stored in a password-protected Excel document. The results are expected to be ready in November 2027.

### Recruitment

Data related to the number of eligible people, approached, recruited, retained for the study (i.e., completion of interview and at least one intervention session), and completion rates of questionnaires will be analysed using descriptive statistics with 95% confidence intervals (CI) (where applicable).

### Demographic and baseline characteristics

Demographic and baseline characteristics will be presented using descriptive statistics such as mean (standard deviation), median (interquartile range), or percentages to represent frequency. The type of descriptive statistic will vary according to the variable type (e.g., continuous, ordinal, dichotomous, etc.). We will present both the overall and within-groups descriptive statistics. We will explore the level of MS severity among participants recruited in the intervention and control group to understand which participant groups were interested in taking part in the feasibility study.

### Feasibility measures

We will use the CONSORT diagram to report data regarding eligible individuals, participants screened, recruited (per month and per site), randomised, and lost to follow-up.

We will report the time to recruit the target sample, possible recruitment issues, the number of participants who book and attend the initial intervention session, the number of sessions booked and cancelled, hours of support received, intervention components delivered, withdrawal and follow-up rates, reasons for withdrawal (if provided), and the number of employers recruited. The hours of intervention received per participant will be used to determine the workforce needed for a future RCT.

Participants who withdraw during the study will be asked (where possible) why they are withdrawing and if they are still interested in completing the end of intervention interview and remaining assessments for the trial.

### Outcome measures

All outcome measures will be summarised using descriptive statistics (as reported for the demographic characteristics) for all time points separately for the intervention group and control.

### Economic evaluation

The cost of delivering MSVR will be estimated from the time MSVR champions spent participating in the training, attending mentoring sessions, and providing the intervention (measured with the session content forms). Data regarding the hourly rate of MSVR champions will be obtained from the MS charities participating in the feasibility trial.

The cost of providing the mentoring will be calculated based on the hourly rate of the mentors and hours of training, mentoring, and administrative support provided.

We will conduct a feasibility economic evaluation to assess whether it is feasible to collect and attribute costs to the resource use data collected.

### Interviews

The interviews will be digitally audio-recorded, transcribed verbatim, and analysed on NVivo v14.0. Interviews will be analysed thematically [43] using a hybrid deductive and inductive approach to develop codes and themes [44]. Thematic analysis involves five steps (familiarisation with the data, generation of initial codes, generating themes, reviewing potential themes, and defining and naming themes) [45]. Any disagreements will be discussed with a third researcher (RdN).

#### Patient and public involvement

A lead PPI representative (White British Man, diagnosed with MS for 25 years, currently not working) has been co-developing this research programme since its inception and will be involved in the study governance and interpretation of the findings. Two further PPI representatives have been recruited to be part of the independent steering committee throughout the trial.

#### Ethics and dissemination

This study has received ethical approval from the School of Medicine Research Ethics Committee (REC) at the University of Nottingham (FMHS 102-0325). This study was registered in ClinicalTrials.gov (NCT06966115).

Any important protocol modifications will be reported to the School of Medicine REC, trial registry, and trial participants when relevant to their participation in the study.

The findings from this feasibility trial will be presented at national and international conferences, published in peer-reviewed journals, and disseminated in webinars and newsletters of the participating charities.

Anonymised data from the trial regarding the intervention delivered and questionnaires will be made available on a repository after the feasibility trial’s completion. Due to ethical considerations, the full transcripts of the end-of- intervention interviews will not be made available; quotes from the interviews will be used to illustrate the themes identified.

## Funding

This study was funded by the UK MS Society (Ref:184). The funder had no role in the design or direction of the project.

## Data Availability Statement

No datasets were generated or analysed during the current study.

## Acknowledgements

The authors would like to thank the patient and public involvement representatives who supported the development of this study.

## Supporting Information

**S1 File. SPIRIT Guidance**

## Notes

### Competing Interest Statement

Author BDP has received funding from the Neurology Academy (speakers bureau) to deliver lectures on vocational rehabilitation for people with MS. Author RdN has received funding (speakers bureau) from Novartis, Biogen, and Merck for delivering lectures on psychological aspects of MS and cognitive screening and rehabilitation in MS.

### Clinical Trial

This study was registered in ClinicalTrials.gov (NCT06966115).

### Author Declarations

This study has received ethical approval from the School of Medicine Research Ethics Committee (REC) at the University of Nottingham (FMHS 102-0325).

## References

[1] Trapp BD, Ransohoff RM, Fisher E, Rudick RA. Neurodegeneration in Multiple Sclerosis: Relationship to Neurological Disability. The Neuroscientist 1999;5:48–57. 10.1177/107385849900500107.

[2] Dobson R, Giovannoni G. Multiple sclerosis – a review. Eur J Neurol 2019;26:27–40. 10.1111/ENE.13819.

[3] Frain MP, Bishop M, Rumrill PD, Chan F, Tansey TN, Strauser D, et al. Multiple Sclerosis and Employment: A Research Review Based on the International Classification of Function. Rehabilitation Research, Policy, and Education 2015;29:153–64. 10.1891/2168-6653.29.2.153.

[4] Strober DL. Determinants of unemployment in multiple sclerosis (MS): The role of disease, person-specific factors, and engagement in positive health-related behaviors. Mult Scler Relat Disord 2020;46. 10.1016/j.msard.2020.102487.

[5] Hennessey ML. Factors Associated with Employment Status. Employment Issues and Multiple Sclerosis. 2nd ed., 2008, p. 19–38.

[6] Smith MM, Arnett PA. Factors related to employment status changes in individuals with multiple sclerosis. Multiple Sclerosis Journal 2005;11:602–9. 10.1191/1352458505ms1204oa.

[7] Teni FS, Machado A, Dervish J, Fink K, Gyllensten H, Friberg E. Factors associated with self-reported work ability among people with multiple sclerosis in Sweden. Mult Scler J Exp Transl Clin 2025;11. 10.1177/20552173241304324/FORMAT/EPUB.

[8] Frank A. Vocational Rehabilitation: Supporting Ill or Disabled Individuals in (to) Work: A UK Perspective. Healthcare 2016;4:46. 10.3390/healthcare4030046.

[9] Doogan C, Playford ED. Supporting work for people with multiple sclerosis. Multiple Sclerosis Journal 2014;20:646–50. 10.1177/1352458514523499.

[10] Leslie M, Kinyanjui B, Bishop M, Rumrill PD, Roessler RT. Patterns in workplace accommodations for people with multiple sclerosis to overcome cognitive and other disease-related limitations. NeuroRehabilitation 2015;37:425–36. 10.3233/NRE-151271.

[11] Machado A, Murley C, Dervish J, Teni FS, Friberg E. Work Adjustments by Types of Occupations Amongst People with Multiple Sclerosis: A Survey Study. J Occup Rehabil 2024;34:461–71. 10.1007/S10926-023-10142-2/TABLES/4.

[12] Bishop M, Park S, Ko E, Koc M, Zhou K, Rumrill P. Employment Accommodation Experiences Among American Workers with Multiple Sclerosis: A Mixed-Method Analysis. J Vocat Rehabil 2025. 10.1177/10522263241310070.

[13] De Dios Pérez B, Das Nair R, Radford K. A mixed-methods feasibility case series of a job retention vocational rehabilitation intervention for people with multiple sclerosis. Disabil Rehabil 2023:1–12. 10.1080/09638288.2023.2181411.

[14] LaRocca NG, Kalb R, Gregg K. A program to facilitate retention of employment among persons with multiple sclerosis. vol. 7. 1996.

[15] De Dios Pérez B, das Nair R, Radford K. Development of a Job Retention Vocational Rehabilitation Intervention for People with Multiple Sclerosis Following the Person-Based Approach. Clin Rehabil 2024. 10.1177/02692155241235956.

[16] De Dios Perez B, Holmes J, Elder T, Lindley R, Evangelou N, das Nair R, et al. Implementing vocational rehabilitation for people with multiple sclerosis in the UK National Health Service: a mixed-methods feasibility study. Disabil Rehabil 2024:1–13. 10.1080/09638288.2024.2417031.

[17] De Dios Perez B, Booth V, das Nair R, Evangelou N, Hassard J, Ford HL, et al. A qualitative study exploring how vocational rehabilitation for people with multiple sclerosis can be integrated within existing healthcare services in the United Kingdom. BMC Health Serv Res 2024;24:995. 10.1186/s12913-024-11424-y.

[18] Bond L, Simmons E, Sabbath EL. Measurement and assessment of fidelity and competence in nonspecialist-delivered, evidence-based behavioral and mental health interventions: A systematic review. SSM Popul Health 2022;19:101249. 10.1016/J.SSMPH.2022.101249.

[19] World Health Organisation. Task Shifting Global Recommendations and Guidelines. Geneva: 2008.

[20] Skivington K, Matthews L, Simpson SA, Craig P, Baird J, Blazeby JM, et al. A new framework for developing and evaluating complex interventions: Update of Medical Research Council guidance. The BMJ 2021;374:n2061. 10.1136/bmj.n2061.

[21] Chan AW, Tetzlaff JM, Altman DG, Laupacis A, Gøtzsche PC, Krleža-Jeriż K, et al. SPIRIT 2013 statement: Defining standard protocol items for clinical trials. Ann Intern Med 2013;158:200–7. 10.7326/0003-4819-158-3-201302050-00583.

[22] Eldridge SM, Chan CL, Campbell MJ, Bond CM, Hopewell S, Thabane L, et al. CONSORT 2010 statement: extension to randomised pilot and feasibility trials. BMJ 2016;355. 10.1136/BMJ.I5239.

[23] Ware JE, Sherbourne CD. The MOS 36-item Short-Form Health Survey (SF-36) I. Conceptual Framework and Item Selection 1992;6:473–83.

[24] Reilly MC, Zbrozek AS, Dukes EM. The Validity and Reproducibility of a Work Productivity and Activity Impairment Instrument. Pharmacoeconomics 1993;4:353– 65. 10.2165/00019053-199304050-00006.

[25] Aarts J, Saddal SRD, Bosmans JE, de Groot V, de Jong BA, Klein M, et al. Don’t be late! Postponing cognitive decline and preventing early unemployment in people with multiple sclerosis: a study protocol. BMC Neurol 2024;24:1–15. 10.1186/S12883-023-03513-Y/FIGURES/1.

[26] Van Der Mei I, Thomas S, Shapland S, Laslett LL, Taylor B V., Huglo A, et al. Protocol for a pragmatic randomised controlled feasibility study of MS WorkSmart: an online intervention for Australians with MS who are employed. BMJ Open 2024;14:e079644. 10.1136/BMJOPEN-2023-079644.

[27] The EuroQol Group. EuroQol - a new facility for the measurement of health-related quality of life. Health Policy (New York) 1990;16:199–208. 10.1016/0168-8510(90)90421-9.

[28] Takasaki H, Chien CW, Johnston V, Treleaven J, Jull G. Validity and reliability of the perceived deficit questionnaire to assess cognitive symptoms in people with chronic whiplash-associated disorders. Arch Phys Med Rehabil 2012;93:1774–81. 10.1016/j.apmr.2012.05.013.

[29] Mapi Research Trust. Perceived Deficit Questionnaire: Scaling and Scoring. 2017. 10.4135/9781412984409.n14.

[30] D’Souza E. Modified Fatigue Impact Scale - 5-item version (MFIS-5). Occup Med (Chic Ill) 2016;66:256–7. 10.1093/occmed/kqv106.

[31] Chen G, Gully SM, Eden D. Validation of a New General Self-Efficacy Scale. Organ Res Methods 2001;4:62–83. 10.1177/109442810141004.

[32] Spitzer RL, Kroenke K, Williams JBW, Löwe B. A brief measure for assessing generalized anxiety disorder: The GAD-7. Arch Intern Med 2006;166:1092–7. 10.1001/archinte.166.10.1092.

[33] Kroenke K, Strine TW, Spitzer RL, Williams JBW, Berry JT, Mokdad AH. The PHQ-8 as a measure of current depression in the general population. J Affect Disord 2009;114:163–73. 10.1016/J.JAD.2008.06.026.

[34] Turner-Stokes L. Goal attainment scaling (GAS) in rehabilitation: a practical guide. Clin Rehabil 2009;23:362–70. 10.1177/0269215508101742.

[35] Watkin C, Phillips J, Radford K. What is a ‘return to work’ following traumatic brain injury? Analysis of work outcomes 12 months post TBI. Brain Inj 2020;34:68–77. 10.1080/02699052.2019.1681512.

[36] Matoliż T, Jurakiż D, Greblo Jurakiż Z, Maršiż T, Pedišiż Ž, Paul Morrey C, et al. Development and validation of the EDUcational Course Assessment TOOLkit (EDUCATOOL)-a 12-item questionnaire for evaluation of training and learning programmes OPEN ACCESS EDITED BY 2023;8:1314584. 10.3389/feduc.2023.1314584.

[37] Avery KNL, Williamson PR, Gamble C, Francischetto EOC, Metcalfe C, Davidson P, et al. Informing efficient randomised controlled trials: exploration of challenges in developing progression criteria for internal pilot studies. BMJ Open 2017;7:e013537. 10.1136/BMJOPEN-2016-013537.

[38] Hooper R. Justifying sample size for a feasibility study. London: n.d.

[39] Julious SA. Sample size of 12 per group rule of thumb for a pilot study. Pharm Stat 2005;4:287–91. 10.1002/pst.185.

[40] Lewis M, Bromley K, Sutton CJ, McCray G, Myers HL, Lancaster GA. Determining sample size for progression criteria for pragmatic pilot RCTs: the hypothesis test strikes back! Pilot Feasibility Stud 2021;7:1–14. 10.1186/S40814-021-00770-X/FIGURES/2.

[41] Moore GF, Audrey S, Barker M, Bond L, Bonell C, Hardeman W, et al. Process evaluation of complex interventions: Medical Research Council guidance. BMJ (Online) 2015;350. 10.1136/bmj.h1258.

[42] Murray E, Treweek S, Pope C, MacFarlane A, Ballini L, Dowrick C, et al. Normalisation process theory: A framework for developing, evaluating and implementing complex interventions. BMC Med 2010;8:1–11. 10.1186/1741-7015-8-63/TABLES/3.

[43] Braun V, Clarke V. Using thematic analysis in psychology. Qual Res Psychol 2006;3:77– 101. 10.1191/1478088706qp063oa.

[44] Fereday J, Muir-Cochrane E. Demonstrating Rigor Using Thematic Analysis: A Hybrid Approach of Inductive and Deductive Coding and Theme Development. Int J Qual Methods 2017;5:80–92. 10.1177/160940690600500107.

[45] Byrne D. A worked example of Braun and Clarke’s approach to reflexive thematic analysis. Qual Quant 2022;56:1391–412. 10.1007/S11135-021-01182-Y/FIGURES/D.

